## supplemental figures for "Investigating the sources of variable impact of pathogenic variants in monogenic metabolic conditions"

Supplemental Figure 1

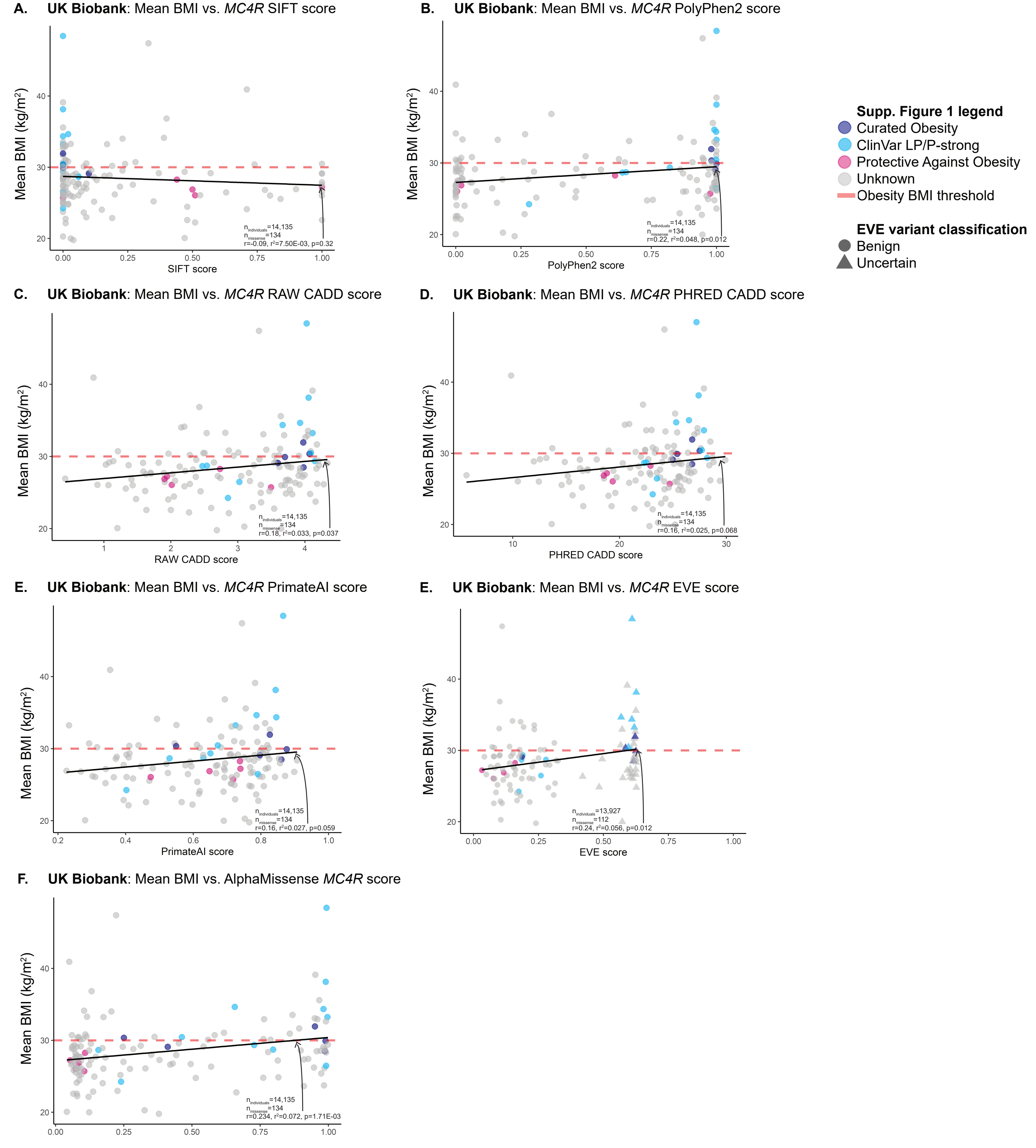


**Supplemental Figure 1: Additional variant pathogenicity predictors do not predict mean phenotype as accurately as ESM1b.** Phenotype correlations were also compared against additional variant pathogenicity prediction methods (**A**-SIFT, **B**-PolyPhen2, **C**-RAW CADD, **D**-PRED CADD, **E**-PrimateAI, **F**-EVE, **G**-AlphaMissense). These methods have lower Pearson correlations with mean BMI compared to ESM1b and do not differentiate between GOF and LOF missense variants in *MC4R*. EVE also does not have full coverage of all *MC4R* missense variants.

Supplemental Figure 2

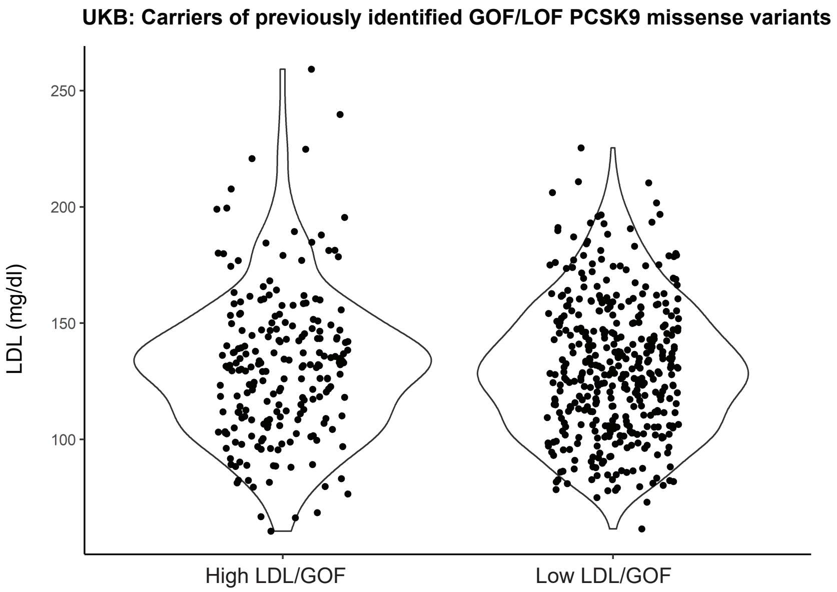


**Supplemental Figure 2: Carriers of GOF/LOF *PCSK9* missense variants do not have significantly different LDL levels in UKB.** Carriers of *PCSK9* GOF (n=216) and LOF (n=398) missense variants were identified. After adjusting for age, sex, and 1st 10 PCs, carrying a GOF or LOF variant was not significantly associated with LDL levels within these carriers.

Supplemental Figure 3

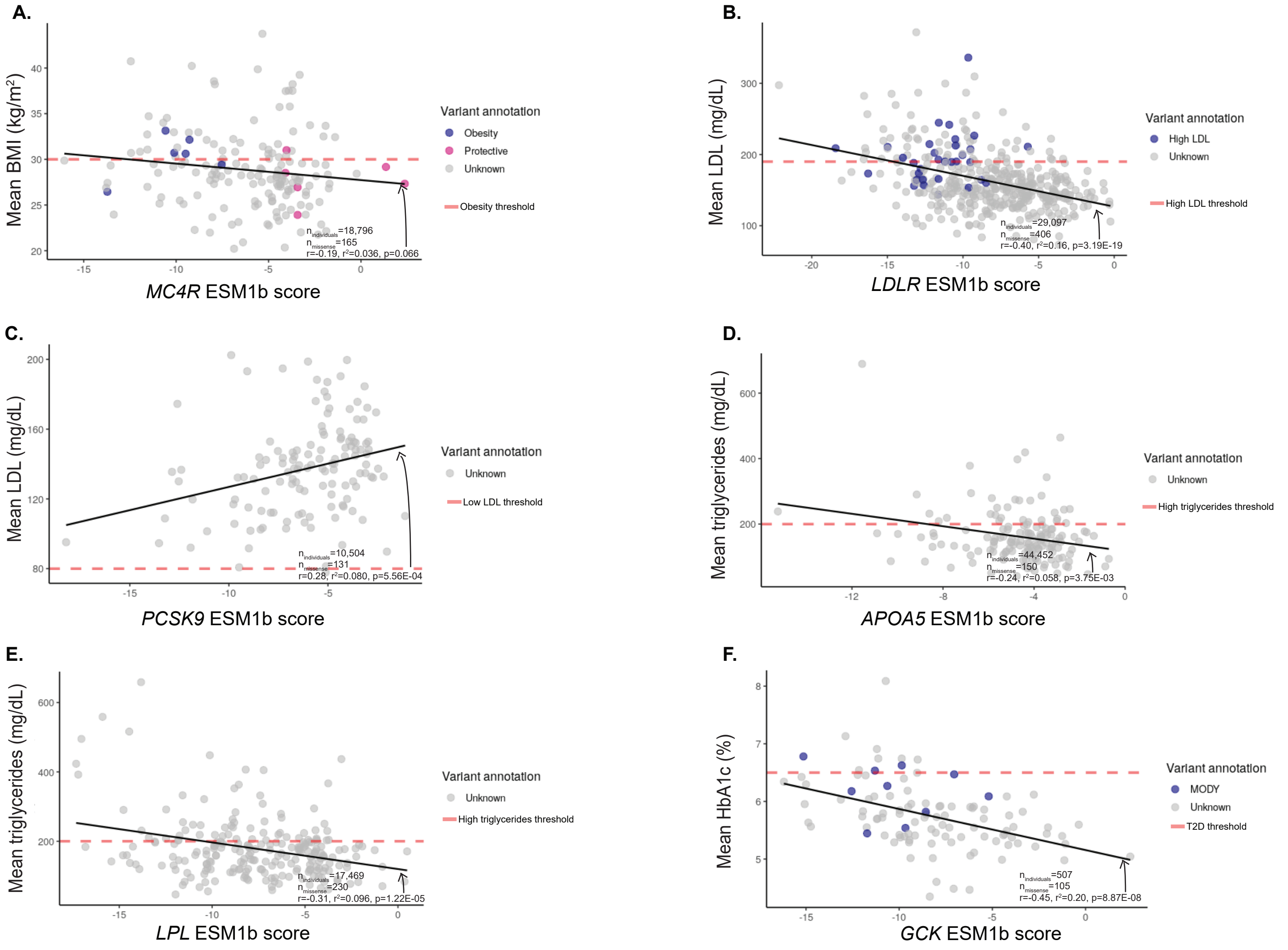


**Supplemental Figure 3: Mean phenotype-ESM1b correlations replicate in UKB 500k exomes.** 5/6 mean phenotype-ESM1b correlations replicate in the 500k exomes: *LDLR* **(B)**, *PCSK9* **(C)**, *APOA5* **(D)**, *LPL* **(E)**, and *GCK* **(F)**. *MC4R* **(A)** correlation approaches significance in this replication. Replication completed in individuals only in the UKB 500k exomes release and not in the 200k exomes release.

Supplemental Figure 4

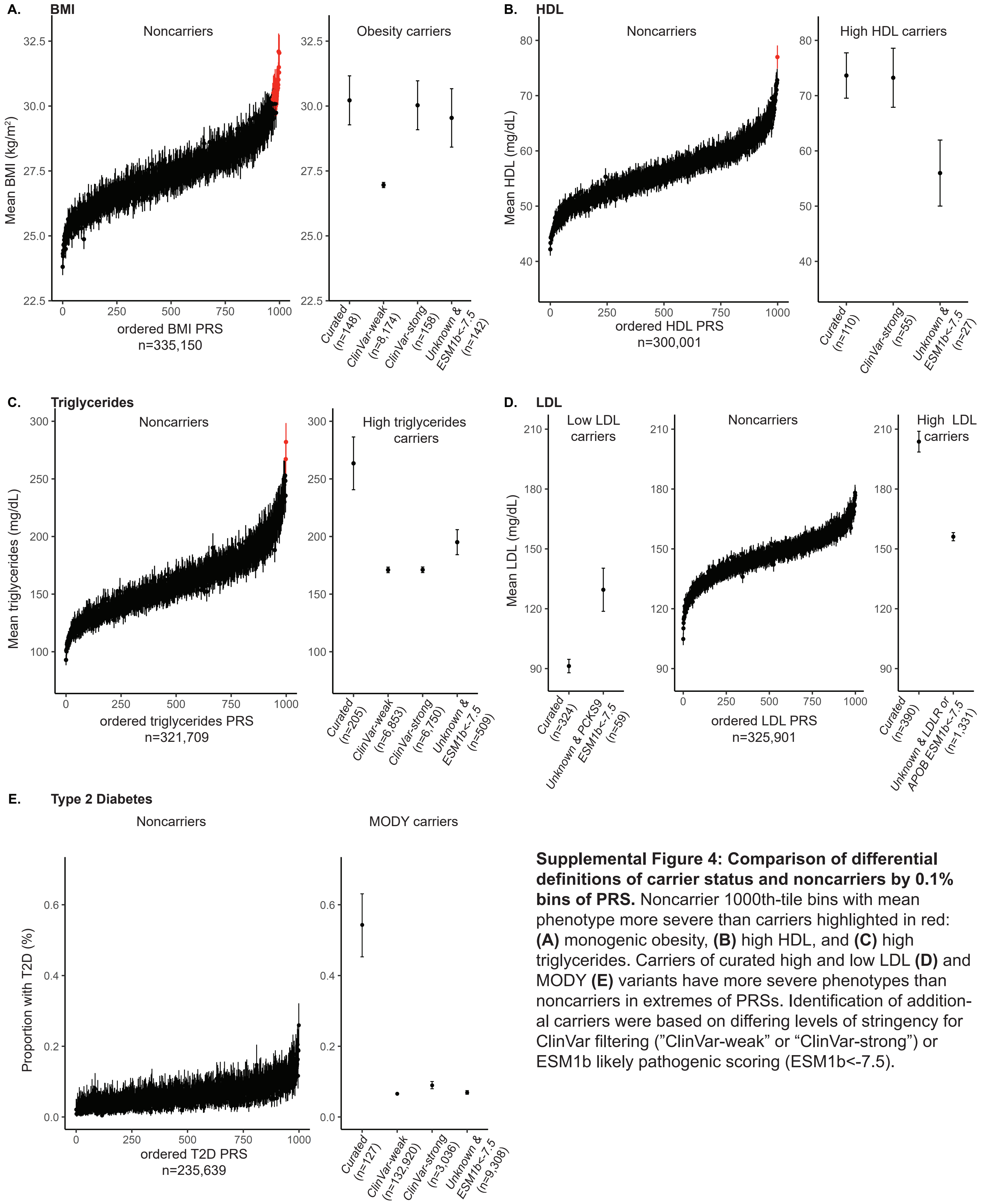


**Supplemental Figure 4: Comparison of differential definitions of carrier status and noncarriers by 0.1% bins of PRS.** Noncarrier 1000th-tile bins with mean phenotype more severe than carriers highlighted in red: **(A)** monogenic obesity, **(B)** high HDL, and **(C)** high triglycerides. Carriers of curated high and low LDL **(D)** and MODY **(E)** variants have more severe phenotypes than noncarriers in extremes of PRSs. Identification of additional carriers were based on differing levels of stringency for ClinVar filtering ("ClinVar-weak" or "ClinVar-strong") or ESM1b likely pathogenic scoring (ESM1b<-7.5).

### Supplemental Figure 5

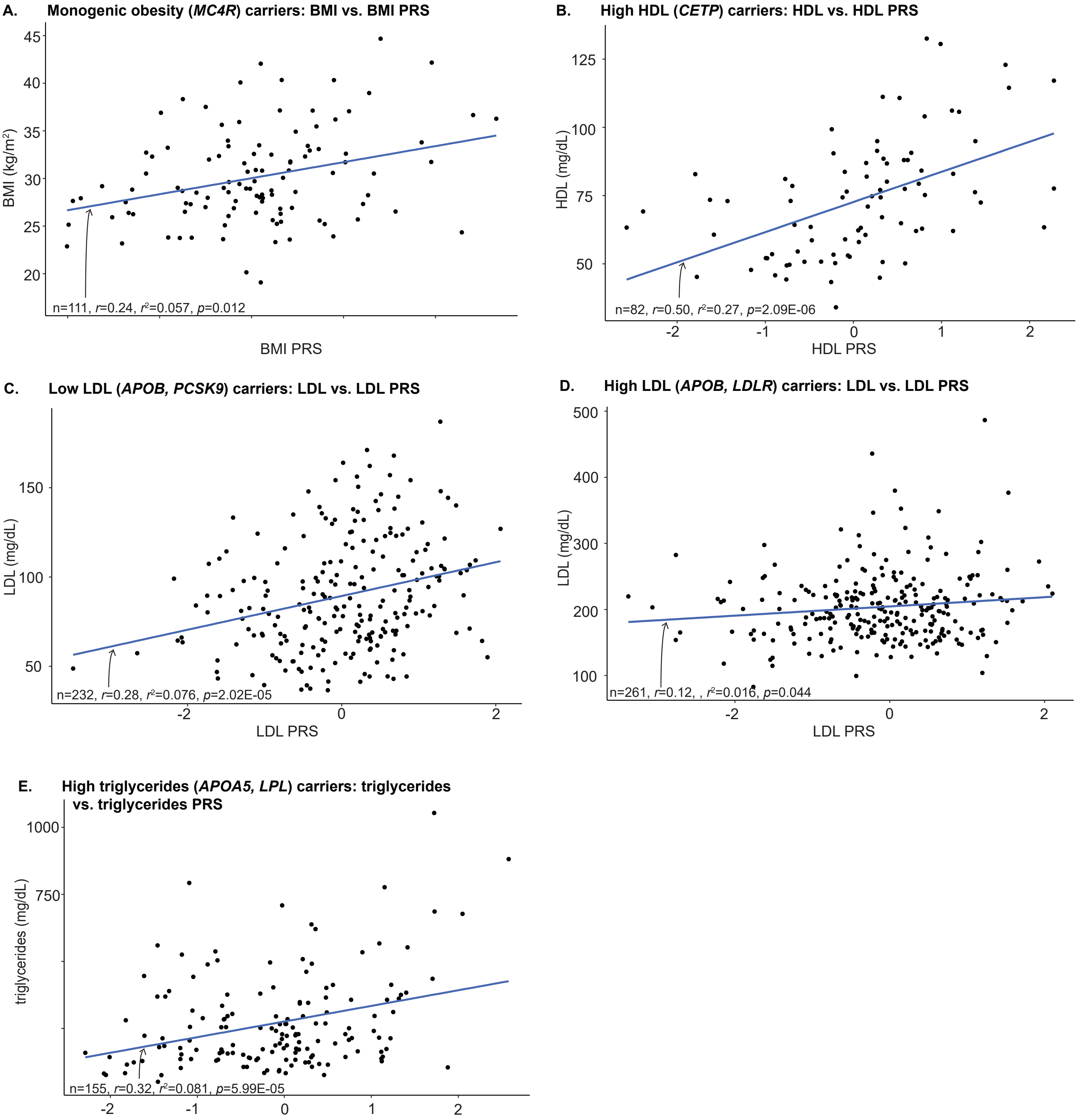


**Supplemental Figure 5: PRS contributes to phenotypic variance even within carriers of pathogenic variants.** After adjusting for sex, age, first 10 genetic PCs, and Bonferroni corrections, corresponding carrier PRS for unrelated, European obesity (A; BMI PRS β=1.68, *p*=5.60E-03), high HDL (B; HDL PRS β=9.79, *p*=1.57E-06), low LDL (β=9.87, *p*=3.18E-06), and high triglycerides (β=62.46, *p*=1.33E-05) carriers were significant. High LDL carriers' corresponding LDL PRS approached significance (β=6.76, *p*=0.028). T2D status of MODY carriers was predicted with T2D PRS after adjusting for age, sex, and first 10 genetic PCs; T2D PRS was not significant in this model (β=0.44, *p*=0.15). Pearson correlation test results are shown on each plot.

Supplemental Figure 6

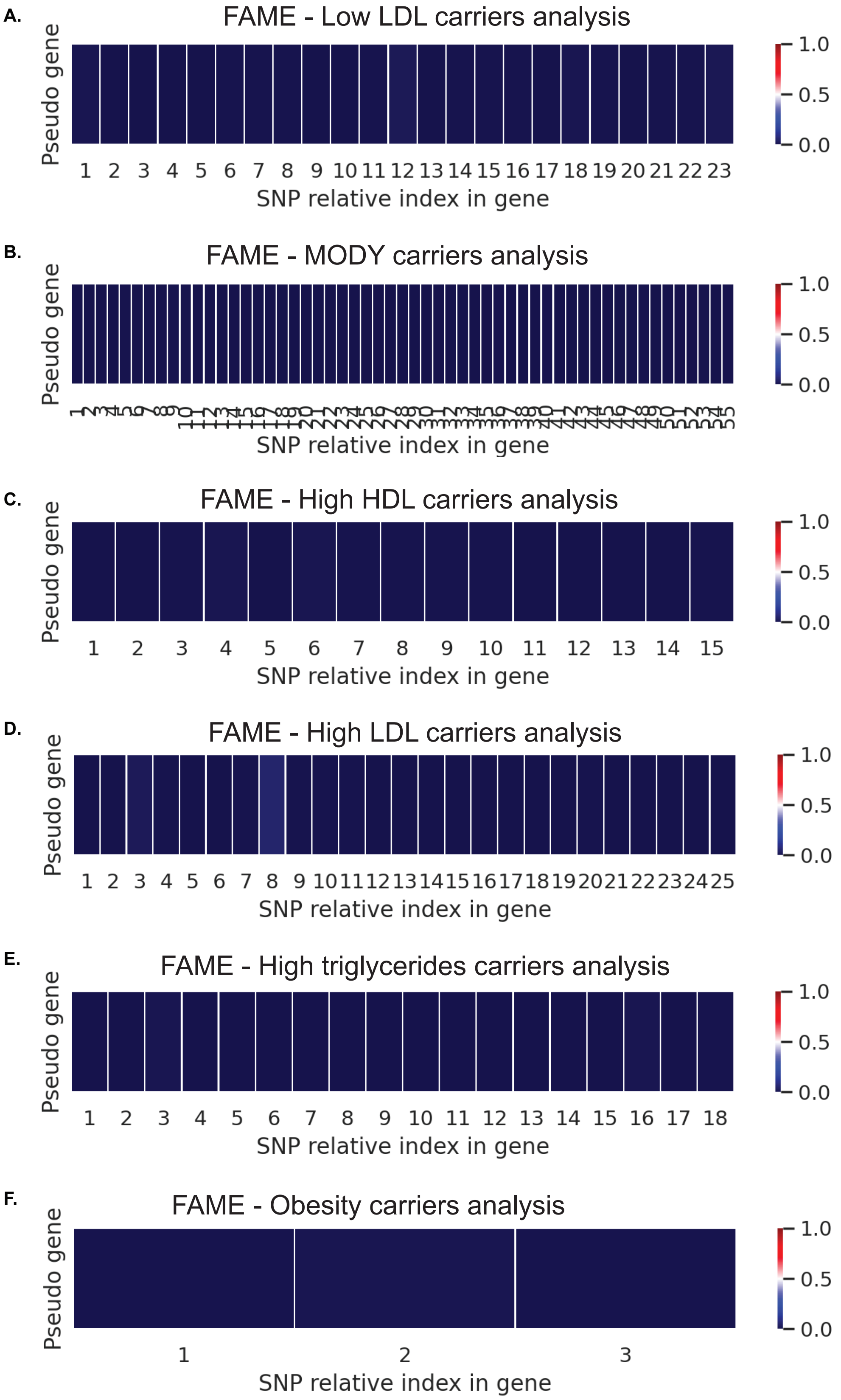


**Supplemental Figure 6: FAME marginal epistasis results are unlikely to be unaffected by LD structure.** Pearson correlations were calculated between pseudo-genes and single nucleotide polymorphisms (SNPs) in the same region of the curated **(A)** low LDL, **(B)** MODY, **(C)** High HDL, **(D)** High LDL, **(E)** high triglycerides, and **(F)** monogenic obesity carriers. Heat maps shown here represent the absolute value of the correlations.
