## supplemental methods for "Investigating the sources of variable impact of pathogenic variants in monogenic metabolic conditions"

Table 1: Pathogenic variant curation methods

| **Pathogenic variant group** | **Identification method** |
| --- | --- |
| Curated pathogenic variants | Variants identified by Goodrich, et al., and Mirshahi, et al., using ACMG/AMP criteria and blinded testing by reviewers |
| ClinVar-weak | Variants in ClinVar with at least one report of “pathogenic” or “likely pathogenic”, but may contain additional conflicting reports |
| ClinVar-strong | Variants in ClinVar with only reports of “pathogenic” or “likely pathogenic” |

Testing for epistasis via PRS

Testing for genetic epistasis occurring between common background genetic variation and monogenic variant carrier status was completed using the model, y=ΣG⋅β_G_+C∙β_C_+ΣG∙C⋅β_CxG_+ϵ, where y is the phenotype of interest, G represents common genetic variation and its associated effect β_G_ on the phenotype of interest, C is an indicator if an individual is carrying a pathogenic variant and β_C_ is the effect size of the monogenic variant on the phenotype of interest, and G∙C is the interaction between common background variation and carrier status (i.e., genetic epistasis) with its associated effect size on the phenotype of interest, β_CxG_. Covariates, such as age, sex, and the individual’s first 10 genetic PCs are also adjusted for in this model. One method this project employed to test for genetic epistasis was to use PRS as a proxy for ΣG⋅β_G_, leading to the model y=β_PRS_⋅PRS+C∙β_C_+C⋅PRS⋅β_CxPRS_+ϵ. Age, sex, and the first 10 genetic PCs were adjusted for in this model.

Additional FAME information

We have defined the target gene(s) proximal region as the specific physical genome region that encompasses the genes we are interested in when analyzing pathogenic variants. By employing this definition, we have divided the *N*✕*M* array genotype matrix (***G***) into two separate matrices: the *N*✕*M_1_* gene-proximal SNP matrix (***G_1_***) containing all the SNPs within the physical coverage of the target genes in array SNP data, of which the pathogenic variants are of interest. For example, the target genes of interest can be *LDLR*, *PCSK9*, *APOA5*, and so on. The remaining *N*✕*M*_2_ gene-distal SNP matrix is ***G_2_***. (We have *M = M_1_ + M_2_*). As a result, our model incorporates the additive effect of both the ***G_1_*** and ***G_2_*** as well as the interactive effect between the pseudo gene of interest (***C_t_***) and the ***G_2_***. The model assumption is formally defined as:

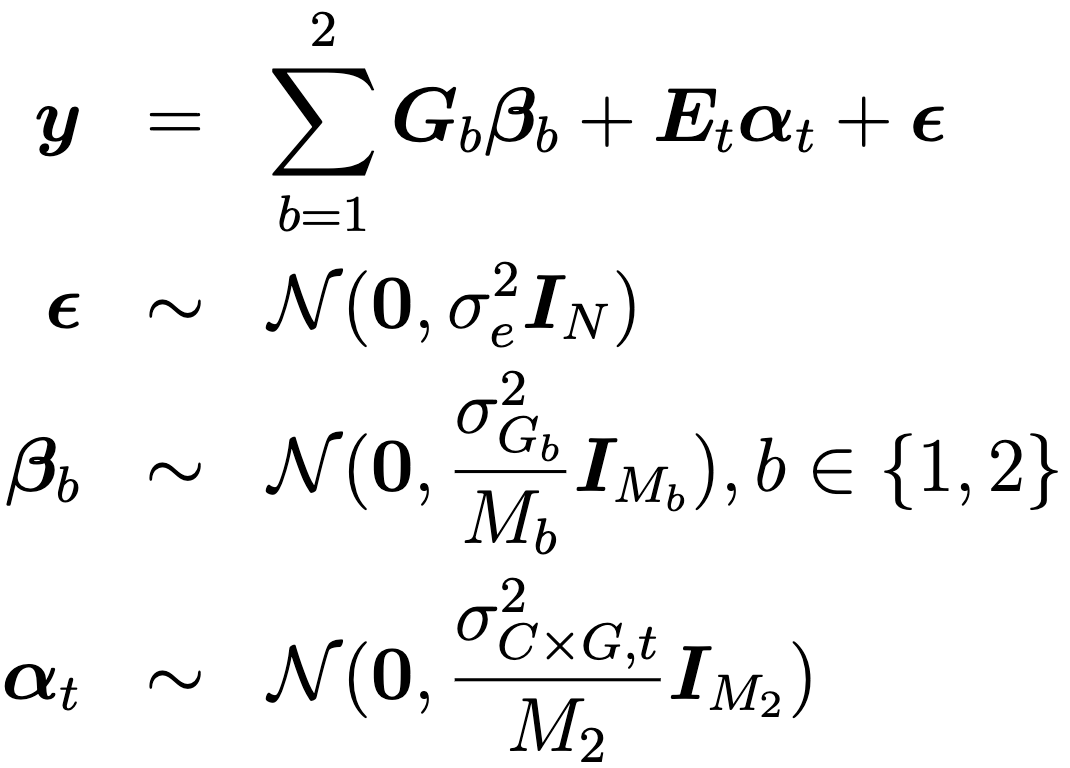

Here
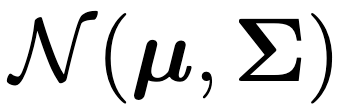
 defines the normal distribution with mean
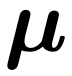
 and covariance matrix
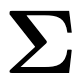
.
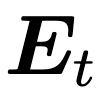
 denotes an *N*✕*M*_2_ pseudo-gene by genetic interaction matrix defined as
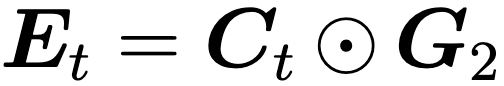
 where
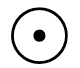
 the row-wise Kronecker product. Here ***C****_t_* represents the target pseudo gene of interest and denotes whether individuals carry a burden of pathogenic variants at the target gene *t* defined as follows: ***C****_t_*=0 if individual *i* does not carry any relevant pathogenic variants at target genes, and ***C****_t_*=1 if individual *i* carries at least one pathogenic variant at the target gene(s) *t*, as detected from the Whole Exome Sequencing (WES) dataset.
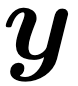
 denotes the residualized phenotypes as an *N*-vector, which was obtained by taking the original phenotype and regressing out the fixed effect of the target pseudo-gene, together with the top 20 PCs, age, and sex. We use the same notation as the previous section, β_C_, to denote the fixed effect of the target pseudo-gene. In this model,
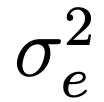
,
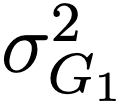
,
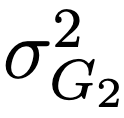
, and
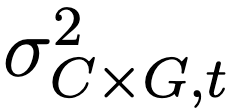
 are the residual variance, additive genetic variance at each bin (
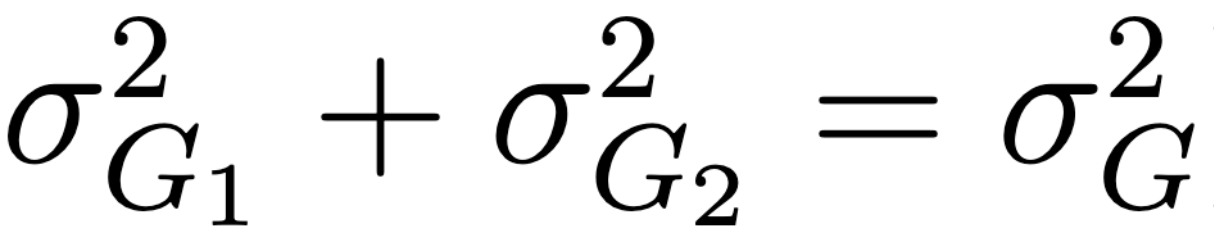
), and the marginal epistasis variance components, respectively.
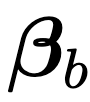
denotes the additive effects of SNPs that are proximal and distal to the target gene(s), while
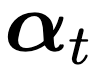
 denotes the interaction effects between target pseudo gene *t* and each of the SNPs in the gene-distal SNP matrix (***G_2_***). Full details can be found in Fu, et al., 2023.^21^
